# Effectiveness of the air-filled technique to reduce the dead space in syringes and needles during ChAdox1-n CoV vaccine administration

**DOI:** 10.1101/2022.04.28.22274427

**Authors:** Patchara Phankavong, Naphatthorn Prueksaanantakal, Nontawat Benjakul, Anan Manomaipiboon

## Abstract

In the current study, we calculated the vaccine volume and amount of dead space in a syringe and needle during ChAdox1-n CoV vaccine administration using the air-filled technique, to reduce dead space in syringe and needle that administer up to 12 doses per vial. The hypothetical situation uses a vial with a similar size as the ChAdox1-n CoV vial. We used distilled water (6.5 ml), to fill the same volume as five vials of ChAdox1-n CoV. When 0.48 ml of distilled water is drawn according to the number on the side of the barrel, an additional 0.10 ml of air can be used in the dead space of the distilled water in the syringe and needle for 60 doses, which can be divided into an average of 0.5 ml dose. ChAdox1-n CoV was administered using a 1-mL syringe and 25G needle to administer ChAdox1-n CoV vaccine into 12 doses using this air-filled technique. The volume of the recipient vaccine will be increase by 20% and save the budget of a low dead space syringe (LDS).

**Graphical Abstract:** 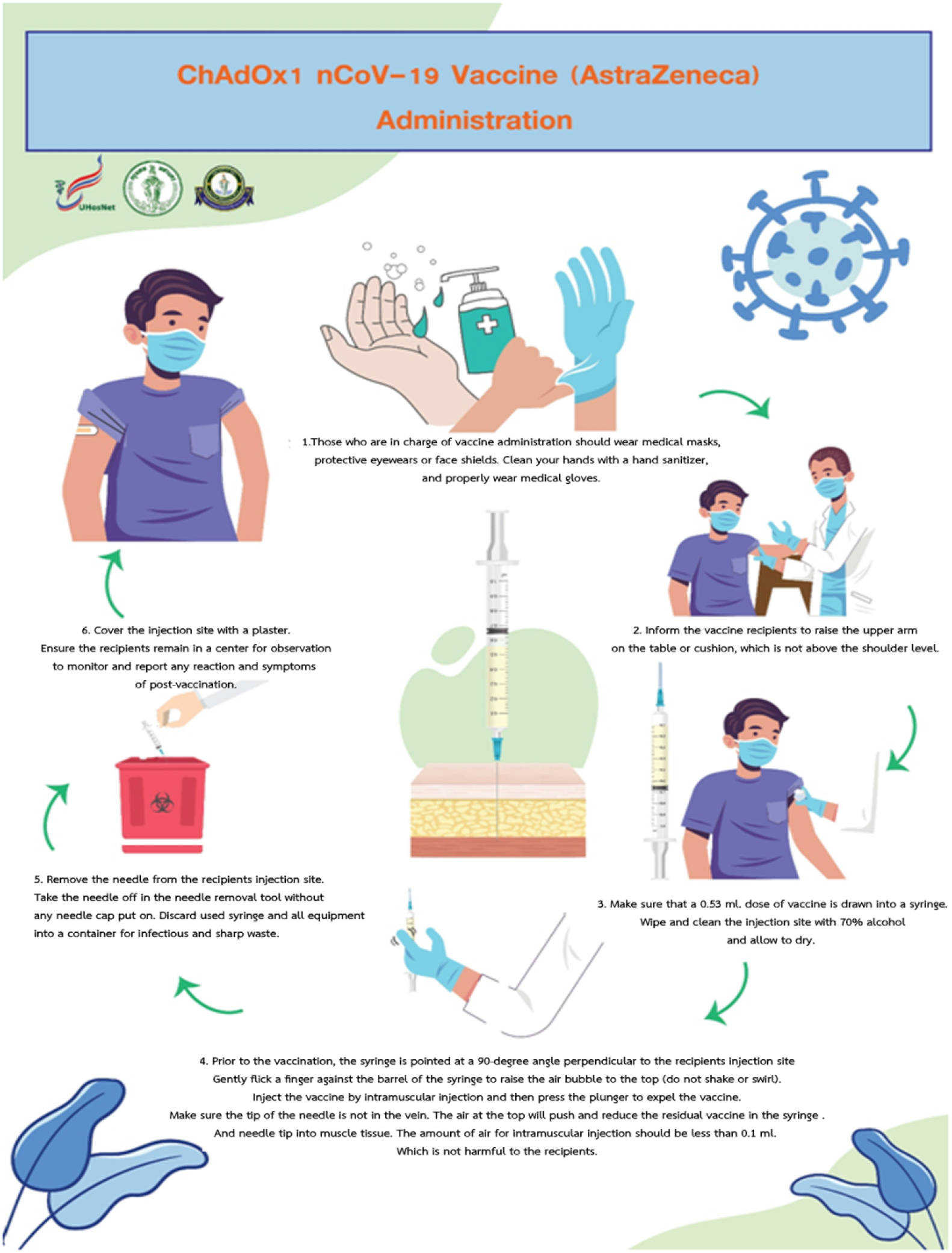

## Introduction

Since December 2019, the SARS-CoV-2 virus epidemic has spread worldwide, leading to the declaration of a pandemic in March 2021[1]. Several countries have endeavored to experiment and create vaccines to prevent and control the transmission of COVID-19 [2]. The main vaccines in Thailand is ChAdox1-n.CoV, which has been approved by the Ministry of Public Health and the Food and Drug Administration. This vaccine was registered on January 20, 2021. However, the limited capacity and vaccination demand of every country has increased beyond supply, creating a shortage of vaccines in most countries, including Thailand.

ChAdox1-n.CoV is used to enhance immunity among people ≥ 18 years of age to prevent symptoms of COVID-19 [3]. This vaccine is a monovalent vaccine consisting of adenovirus from chimpanzees (ChAdOx1) encrypted by the glycoprotein S of SARS-CoV-2 [4]. After injection, the glycoprotein S of SARS-CoV-2 strengthens the immune system and prepares them for later infection.

A vial of ChAdox1-n.CoV can be divided into ten doses (multiple doses) of 5 ml for ten people. The manufacturers generally overfill the vial to replace the dead space in the syringe and needle in the preparation and injection procedure for ten people. The amount of overfilled vaccine is the same in every brand for multiple dose vials, including Pfizer, which has sufficient vaccines for six people. Therefore, in many countries that have reserved COVID-19 vaccines, the multiple dose type is assigned to use an low dead space (LDS) syringes and needles to reduce the wasted volume that remains in the tip of the syringes and needles [5]. However, the LDS and needle cost is 4 to 10 times more than a 1-ml syringe and 25G needle (1 to 1.5-inch length).Therefore the latter syringes and needle needles are commonly used. Additionally, no manufacturers for LDS and needles are available in Thailand, and they need to be imported from overseas. Moreover, there is a worldwide shortage of vaccines. Therefore, a study about vaccine administration, including the preparation and injection procedure of ChAdox1-n.CoV, to allow the vaccine to be divided to 12 doses per vial instead of 10, as limited by the manufacturer, is required. This procedure will help clinics suitably administer the vaccine in the most efficient manner, which will mitigate the supply issues and unavailability of LDS needles in many countries [6].

## Materials and Methods

This research was approved by the Ethics Committee at our hospital, registration no. 096/2564.All procedures were performed after written informed consent from all participants.

The research procedure was as follows:

- The test was first performed using distilled water to test the hypothetical situation.
- A vial with a similar size as the ChAdox1-n CoV vial was used. Distilled water of the same volume as a single vial of ChAdox1-n CoV(6.5 ml) was filled in the vial.
- A digital precision balance weighing scale was prepared to measure empty syringes and vaccine (in mg) up to four decimal places and pass the calibration of National Institute of Metrology Thailand (NIMT) as described below.

Step 1: 60 sets of 1-ml syringe and 25G needle were weighed and recorded, with a length of 1 inch needle, and the weighted average was calculated.

Step 2: 0.45 ml water with 0.1 ml air was aspirated, resulting in < 0.5 ml water being ejected from the syringe.

Step 3: 0.48 ml of distilled water and an additional 0.10 ml of air was drawn into the syringe to reduce the dead space in the syringe and needle. The amount of the water appeared to be slightly more than 0.5 ml. Thereafter, the weight of 60 sets of syringe and needle were recorded and weighted average was calculated.

Step 4: 0.5 ml of distilled water from the barrel was discarded, syringe and needle were weighed, the weight recorded,, and the weighted average calculated.

- The experiment using the same procedures was repeated until 12 doses of distilled water were obtained per vial, for which the amount in every dose was ≥ 0.5 ml.
- The second stage involved using ChAdox1-n CoV in the real situation in the same way as in the hypothetical situation as described below.

Step 1: Sixty sets of 1-ml syringes and 25G needles, with a length of 1 inch needle, were weighed, the weight recorded, and the weighted average calculated. Five vials of vaccine were used. Because a vial can be divided into 12 doses, five vials would comprise 60 doses.

Step 2: The vaccine was prepared by stabbing the needle in the vaccine vial and drawing in 0.48 ml of vaccine and an additional 0.10 ml of air to reduce the dead space in the syringe and needle. The amount of the vaccine would appear to be equal to or slightly more than 0.5 ml. Thereafter, the 60 syringe and needle sets were weighed, the weight recorded, and the weighted average calculated.

Step 3: The vaccine was injected to the muscle of the vaccine recipient following the vaccine administration instructions composed by the Nursing Department of the Faculty of Medicine, Vajira Hospital, Navamindradhiraj University. Thereafter, the 60 syringe and needle sets were weighed, the weight recorded, and the weighted average calculated.

### ChAdox1-n CoV Injection Technique

The vaccine injection using the same needles to draw vaccine from the same vial could allow the diseases to spread easily, such as Hepatitis B, Hepatitis C, and human immunodeficiency virus (HIV). Thus, the hospital and universal infectious control guidance precautions were strictly adhered to the aseptic technique. The vaccine injection procedures are described below.

1. Medical personnel must wear a surgical mask, glasses, and face shield, and sanitize their hands with hand sanitizer and wear suitable gloves.
2. The medical personnel and recipients receiving injection must be explained to put one of their upper arms on the table or on the surface, which is not higher than shoulder level, and not flex their shoulder and upper arm muscles during the injection.
3. The syringe should be checked if it contains is 0.5 ml of the vaccine as prescribed by Astra Zeneca. The skin where the vaccine is to be injected is cleaned with 70% alcohol and left to dry.
4. Before injecting the vaccine, the syringe should be pointed at 90 degrees to let the air flow to the top of the barrel. Flicking the finger lightly on the barrel helps the air bubble move to the top (the syringe should not be shaken). Next, needle should be inserted through the subcutaneous layer to the muscle ensuring that the tip of the needle is not in the blood vessel, and inject the vaccine by pushing the plunger to the end. The air on the top part of the syringe pushes the dead space in the syringe and needle to the muscle by a slight amount. Therefore, less than 1 mL air might pass to the muscle, which does not cause any harm. If the medical personnel and recipients receiving injection have a thick subcutaneous layer, a 1.5-length 25G needle should be used instead.
5. the needle should be pulled out from the vaccine recipient and removed in the needle clipping devices without using the luer lock. The syringe and all other equipment used should be discarded in the infectious waste disposal container.
6. Finally a band-aid should be placed on the injection wound and the vaccine recipient should be asked to wait in the waiting area to observe whether any symptoms occur after the injection.

The researcher should check the specific gravity in the five vials of the ChAdox1-n CoV used in the experiment to calculate the volume that the patient has received.

### Statistical Analysis

The data was analyzed using SPSS Program, Version 28 (Chulalongkorn University Bangkok Thailand) to calculate the weighted average of the syringe, needle, vaccine, volume, and standard variation.

## Results

In the experiment using 6.5 ml of the distilled water in the vial, having the same size as the ChAdox1-n CoV vial, 0.48 ml of distilled water and an additional 0.1 ml of air to replace the dead space of the distilled water in the syringe and needle was drawn. The distilled water was administered to 12 doses per vial. The weighted average in the syringe before injecting was evaluated to be 0.5168 ml, the weighted average of dead space 0.0122 ml, and the volume of distilled water injected out of the syringe 0.5046 ml.

The ChAdox1-n CoV vaccine administration experiment used a normal syringe and needle to draw in 0.48 ml of the vaccine, according to the number on the side of the barrel and an additional 0.10 ml of air to reduce the dead space in the syringe and needle. We then used this procedure to prepare 0.5 mL or slightly more of ChAdox1-n CoV vaccine using a 1-ml syringe and 25G needle to draw in the 0.48 ml of the vaccine, according to the number on the side of the barrel, and an additional 0.10 ml of air to reduce the dead space in the syringe and needle x. We could administer 12 doses per vial. The weighted average of the vaccine before injecting was found to be 0.5048 ml and the weighted average of the dead space 0.0263 ml. Thus, the volume of the vaccine that the recipient would receive was 0.5048 ml (the weight of the vaccine that the recipient would receive was = 0.5199/1.030 [sp.gr. of the vaccine])

## Discussion

This experiment was aligned with research on antibody level for SARS-CoV-2 after injection of the 1^st^ dose of ChAdox1-n CoV-19 using 12 doses per vial [8]. We found that approximately 8.57 weeks after injecting 12 doses per vial, the level of immunity was higher at a positive value of 93.3%, and higher among women than men. Additionally, the immunity did not vary according to age, disease, body mass index, blood type, and blood pressure, and corresponded with the advice from the Pfizer-BioNTech COVID-19 Vaccine, which uses LDS to divide six doses of vaccine per vial. After injecting the vaccine to the recipient, we found ≤ 0.035 ml dead space in the syringe. From this experiment using the air-filled technique to reduce the dead space in the syringe and needle, 0.0263 ml of the vaccine remained. Thus, the vaccine recipient will receive 0.5048 mL volume of the vaccine using the 1-ml syringe and 25G needle instead of the LDS. This technique will reduce the expenses of importing LDS, which costs 2.78 THB per syringe compared to 1.65 THB for a 1 ml syringe and 0.43 THB for a 25G needle. This administrative technique increases the amount of the recipient vaccine by 20% and is more economized than using LDS. This will help the Nursing Department of the Faculty of Medicine, Vajira Hospital to create guidelines and strategies to administer the vaccine using the air-filled technique to reduce the dead space in the syringe and needle, as well as to publish an interdisciplinary method in vaccine administration for all vaccine recipients.

## Conclusion

ChAdox1-n CoV vaccine administration using a 1-ml syringe and 25G needle to draw in 0.48 ml of vaccine, according to the number on the side of the barrel, and an additional 0.10 ml air, to reduce the dead space in the syringe and needle, to achieve an average volume of 0.5 ml (or slightly more), allowed the vaccine to be administered at 12 doses per vial. The air-filled and vertical injection techniques included flicking the finger lightly on the barrel to let the air flow to the top of the barrel. Then, the syringe was pointed at 90 degrees to pierce the muscle before pushing the plunger to the end. The air on the top pushed the dead space at the tip of the syringe and needle. We found that 0.0263 ml of dead space remained.

## Data Availability

All data used in the preparation of the manuscript are related to the previously published article and can be found in the Mendeley Data repository: ‘ChAdOx1nCoV-19 vaccination,’ Mendeley Data, V1, doi: 10.17632/yjhssw33cw.1

## Acknowledgment

We wish to thank Assoc. Prof Anayn Manomaipiboon, the chancellor of Navamindradhiraj University for providing advice and ideas relating to all aspects of this research.

We wish to thank Dr. Nontawat Benjakul, MD, Faculty of Medicine, Vajira Hospital in Anatomical Pathology, Navamindradhiraj University for his assistance with the writing of the manuscript and for proofreading the final document.

We wish to thank Ms. Yupin Chalermchai, Ms. Pensiri Kaewkasikarn, Ms. Chantana Jinawong, Ms. Warisa Jirawatin, and all the registered nurses and staff for their support.

## Author contributions

AM conceptualized the method, designed experimental protocols and validation tests; PP and NP conducted the experiments; AM and PP conducted data analysis and visualization; and PP and NB wrote the manuscript.

## Ethics committee approval

The experiment was performed in accordance with the principles of the Declaration of Helsinki and Good Clinical Practice. This study was approved by the Vajira Institutional Review Board, Faculty of Medicine, Vajira Hospital, Navamindradhiraj University. The vaccine used was authorized by the Thai FDA and the Department of Medical Science (DMS).

## Conflict of Interest Statement

All the authors declare that they have no competing interests.

## Funding source

This work was supported by a grant from Navamindradhiraj University

## Data availability statement

All data used in the preparation of the manuscript are related to the previously published article and can be found in the Mendeley Data repository: “ChAdOx1nCoV-19 vaccination,” Mendeley Data, V1, doi: 10.17632/yjhssw33cw.1

## Notes

### Clinical Trial

NCT04961385

